# Autoantibody-Driven Monocyte Dysfunction in Post-COVID Syndrome with Myalgic Encephalomyelitis/Chronic Fatigue Syndrome

**DOI:** 10.1101/2025.01.09.25320264

**Authors:** Alexander Hackel, Franziska Sotzny, Elise Mennenga, Harald Heidecke, Kai Schulze-Foster, Konstantinos Fourlakis, Susanne Lüders, Hanna Grasshoff, Kerstin Rubarth, Frank Konietschke, Tanja Lange, Carmen Scheibenbogen, Reza Akbarzadeh, Gabriela Riemekasten

## Abstract

Post-COVID syndrome (PCS) has emerged as a significant health concern with persisting symptoms. A subset of PCS patients develops severe myalgic encephalomyelitis/chronic fatigue syndrome (pcME/CFS). Dysregulated autoantibodies (AABs) have been implicated in PCS, contributing to immune dysregulation, impairment of autonomous nerve and vascular function. As recently shown in autoimmune diseases, IgG fractions translate disease-specific pathways into various cells. Therefore, we asked whether IgG fractions from PCS patients could be applied in-vitro to identify specific cytokine responses for PCS patients without (nPCS) and with pcME/CSF. To assess this, we have stimulated monocyte cell lines with IgG fractions from PCS patients. Our findings reveal distinct cytokine responses induced by patient derived AABs which are suggested in vascular and immune dysfunction. In contrast to nPCS, pcME/CSF AABs induced enhanced neurotrophic responses, characterized by significant cytokine correlations involving brain-derived neurotrophic factor (BDNF), glial cell-derived neurotrophic factor (GDNF) and tumor necrosis factor superfamily member 14 (LIGHT). Further, AAB-induced cytokine levels correlate with clinical symptoms. This study emphasizes a contribution of AABs in PCS, in mitigating long-term immune dysregulation, and a need for therapies modulating IgG-induced signaling pathways.

## Introduction

Post-COVID syndrome (PCS) has become a major public health concern, affecting 10–20% of individuals after SARS-CoV-2 infection. Symptoms persist for a minimum of two months and often severely impair daily functioning. [1–3] Among these patients, a subgroup fulfills the Canadian Consensus Criteria (CCC) for diagnosis of myalgic encephalomyelitis/chronic fatigue syndrome (ME/CFS) [4–6]. The clinical overlap between PCS and ME/CFS highlights the shared but poorly understood mechanisms underlying these conditions. Emerging evidence implicates a complex interplay of immune dysregulation, endothelial dysfunction, and vascular anomalies in PCS, which may differ from these in pcME/CFS [7–9]. Functional autoantibodies (AABs) have been identified as significant contributors to immune dysregulation in both acute and post-viral syndromes [10]. For example, AABs against renin-angiotensin system (RAS)-related proteins, including angiotensin-converting enzyme 2 (ACE2) and angiotensin type-1 receptor (AT1R), correlate with disease severity in acute COVID-19 and vascular dysfunction in PCS [11]. Our studies suggest a contribution of specific AABs in inflammatory conditions. As an example, AT1R AABs induced skin and lung inflammation as well as endothelial apoptosis [12, 13]. In ME/CFS, disease-specific alterations in the G-protein coupled receptor (GPCR) AAB network have been associated with key symptoms such as fatigue, muscle pain, and neurocognitive impairments [14, 15]. Similar disruptions in PCS suggest a shared mechanism of immune-mediated dysfunction, with GPCR AABs potentially modulating both inflammatory and neurotrophic pathogenic alterations. Intriguingly, in PCS, AABs against adrenergic receptors were identified as key markers distinguishing symptom severity, emphasizing their contribution to vascular dysfunction and endothelial impairment [16]. Similarly, in PCS, AABs targeting vasoregulatory and immune-modulatory proteins have been linked to chronic vascular inflammation and immune dysregulation.

Monocytes, as pivotal responders of innate immunity, are emerging as key mediators in the immunopathogenesis of PCS and pcME/CFS. In viral infections, monocytes act as first-line effectors, contributing to immune defense and tissue repair. However, their dysregulation can lead to pathological inflammation and tissue damage, as seen in macrophage activation syndrome during severe COVID-19 [17, 18]. Monocyte-driven responses, particularly through secreted cytokines such as CCL18, are also implicated in fibrotic and neurodegenerative processes characteristic for autoimmune diseases such as systemic sclerosis, but also for PCS [19]. As shown by our group, IgG fractions from patients with autoimmune diseases induce a disease-specific protein pattern and pathways in monocytes [12]. The IgG-induced secretome could thus serve as a biomarker for pathways present in their corresponding donor and as a diagnostic tool. To address this hypothesis, the current study investigates the role of AAB-induced monocyte activation in PCS and pcME/CFS. By focusing on the monocyte secretome, this research aims to elucidate whether known mechanisms in ME/CFS such as inflammatory and neurodegenerative processes are induced by AABs, whether IgG-induced cytokines and proteins show associations with clinical findings, and whether the cytokine signature could be used as diagnostic tool. Our study represents the first comprehensive investigation into the interaction between AABs, monocyte activation, and disease pathophysiology in PCS and pcME/CFS, offering new insights into potential therapeutic targets.

## Methods

### Cohort characteristics

Female participants between 24 and 51 years were recruited at the Charité Fatigue Centre, Berlin. Serum samples were collected from 24 PCS patients with persistent fatigue and exercise intolerance following mild to moderate acute SARS-CoV-2 infection and from 12 age-matched healthy controls (HCs). 12 out of 24 PCS patients fulfilled the Canadian Consensus Criteria (CCC) for diagnosis of ME/CFS (pcME/CFS), others were categorized as PCS none ME/CFS (nPCS), the PCS group includes both defined Subgroups. Most patients had a disease duration >6 months, in two of 11 pcME/CFS patients, and in two of 13 PCS patients with a disease duration < 6 months, the diagnosis was confirmed at month 6.

Detailed cohort information is displayed in **Table 1**. Disease and symptom severity were assessed by questionnaires: The functional disability was evaluated by Bell score, ranging from 0 to 100 (with 100 for no restrictions) [20]. PEM severity was evaluated according to Cotler et al., with scores ranging from 0 to 46 (no to frequent/severe PEM) [21]. The severity of the key symptoms, fatigue, pain, and cognitive impairment was quantified using a Likert scale (1 = no symptoms to 10 = severe symptoms). Fatigue was additionally evaluated using the Chalder Fatigue Scale from 0 to 33 (no to heavy fatigue) [22]. Autonomic dysfunction was assessed using the Composite Autonomic Symptom Score 31 (COMPASS-31), ranging from 0 to 100 (no to strongest impairment) [23]. Clinical data was collected and managed using REDCap electronic data capture tools hosted at Charité University Medicine Berlin [24, 25]. Whole blood samples from participants were allowed to clot at room temperature and then centrifuged at 2000 x g for 15 min at 4°C. The purified serum was stored at −80°C for further analysis. SARS-CoV-2 Serology was performed at the Institute for Immunology at Charité Universitätsmedizin Berlin using Anti-SARS-CoV-2 ELISA (IgG) according to manufacturer protocol (Euroimmun Medizinische Labordiagnostika AG, Lübeck, Germany). This study was approved by the Ethics Committee of Charité Universitätsmedizin Berlin (EA2/067/20, EA2/066/20). The participants gave written informed consent.

**Table 1.**
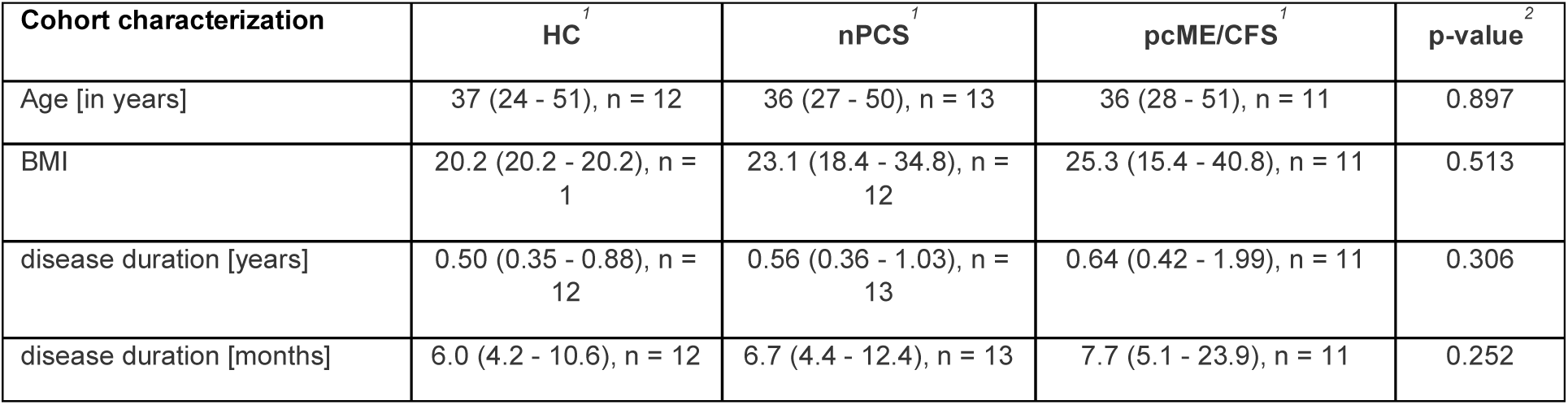
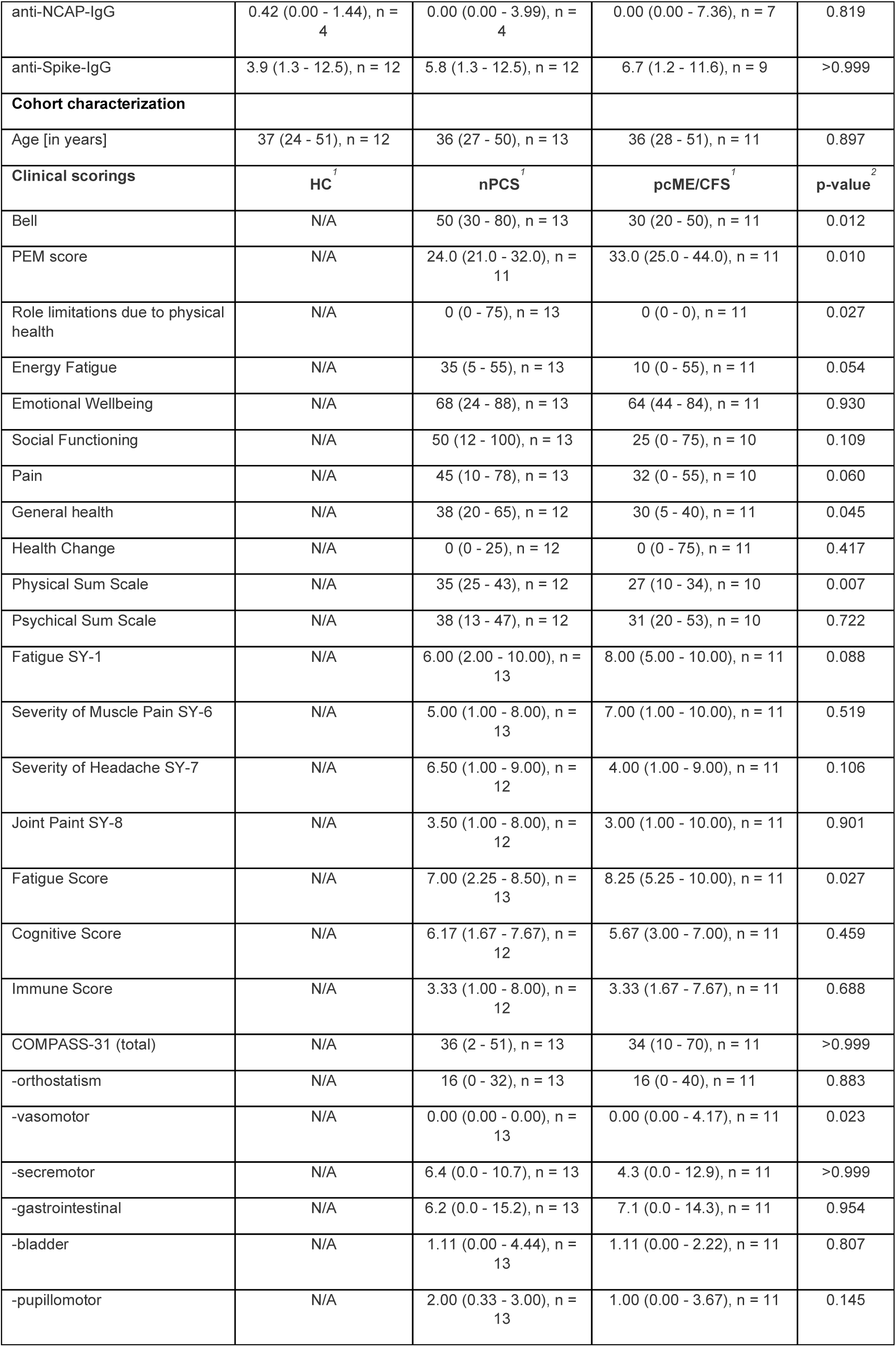
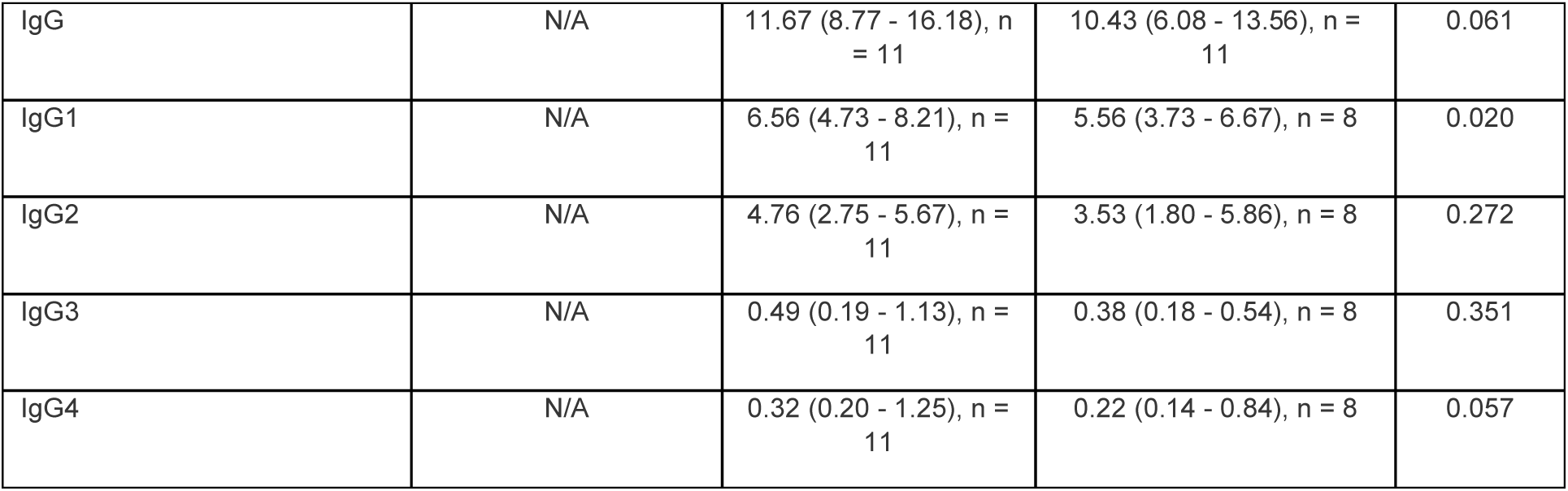
Clinical scorings and cohort characterization. Continuous variables were expressed as median and range. Univariate comparisons of study groups were done using the Kruskal– Wallis test (three groups) or the Mann-Whitney-U-Test (two groups). A two-tailed p-value of <0.01 was considered statistically significant. [n. a. = not assessed].

### Culturing and Preparation of Cells

Human monocytic U937 cells (ACC-5, DSMZ-German Collection of Microorganisms and Cell Cultures GmbH) were cultured in VLE RPMI 1640 medium (Bio & Sell, #F1415) supplemented with 10% heat-inactivated fetal calf serum (Bio & Sell #FCS-ULE1415), 100 U/ml penicillin, and 100 µg/ml streptomycin (BioWest #L0022). The cells were incubated under sterile conditions at 37 °C with 5% carbon dioxide in culture flasks. For stimulation experiments, U937 cells were seeded at a density of 2 × 10^6^ cells/ml one hour before stimulation in 24-well plates (Sarstedt, #83.3922500).

### Isolation of Human IgG

Human IgG was isolated from the patient serum using affinity chromatography at room temperature. Serum samples were diluted fourfold in binding buffer (20 mM Na2HPO4+20 mM NaH2PO4, pH 7) and centrifuged at 10,000 x g for 10 minutes at 4 °C. The supernatant was aspirated from below the lipid layer and transferred to a fresh tube. HiTrap protein G Sepharose columns (Cytiva, Sweden, #17040401) were equilibrated with binding buffer, and the diluted serum was passed through the columns. Unbound molecules were washed out, and bound IgG was eluted using 100 mM glycine (pH 2.7). The eluate was immediately neutralized with 1 M TRIS buffer, then diluted with PBS (BioWest, #L0615) to 15 ml and passed through centrifugal filter units (Amicon, #UFC901024) at 3,000 g and 4 °C. After replenishing and re-centrifuging, the retentate, the IgG concentration was measured using Bradford’s reagent (BioRad #5000006) according to the manufacturer’s instructions.

### Stimulation of U937 Cells with IgG from Patients and Healthy Controls

For stimulation experiments, U937 cells were exposed to purified IgG from three groups: a) nPCS, b) pcME/CFS, and c) COVID-recovered HC. 6.67 μM IgG preparation was added to the cells [13]. After stimulation for 24h, the cells were centrifuged, and the supernatants were stored at −20 °C for cytokine analysis. The levels of cytokines in the supernatants were analyzed using cytokine antibody array glass chips (RayBiotech, Norcross, GA, USA, #AAH-CYT-G5-8). The array protocol was followed as per the manufacturer’s instructions: Non-specific binding sites on the chamber slides were blocked using a blocking buffer. Following this, the supernatants were incubated on the slides with a biotinylated antibody cocktail. After multiple washes, streptavidin-fluorochrome reagent HiLyte Plus™ Fluor 555 was added. The slides were then laser-scanned at 532 nm using an Innoscan Microarray Scanner (Innopsys, France). Background signals were subtracted, and the signals were normalized to positive controls before further analysis using Agilent Scan Control Software ver9.1.

### Data Preparation

Data preparation was performed using the array manufacturer’s batch correction tool available at RayBiotech’s online platform. Raw signals were first corrected for background by subtracting the median background intensities. Subsequently, signal normalization was performed. To normalize distribution and stabilize variance, the data underwent log2 transformation. Further processing of the data was performed using the limma package in R software to eliminate non-biological variance between different experimental batches. This additional step enhanced the reliability of subsequent statistical analyses, enabling the detection of meaningful biological signals amidst technical variability.

### Statistical Analysis

Descriptive Statistics were conducted to characterize the study groups, using medians and ranges for numerical data due to the small sample size. For exploratory univariate analyses, non-parametric tests were applied, including Kruskal-Wallis tests and Mann-Whitney *U* tests. Principal Component Analysis (PCA) was performed on the batch-corrected dataset to explore variance structure and clustering among the PCS, pcME/CFS, nPCS, and HC groups. PCA plots provided a visual representation of the separation between groups, highlighting clustering patterns. PCA visualizations were generated using the ggplot2 package in R. Linear Modeling was conducted to identify cytokines with differential expression between groups using the limma package. Comparisons included PCS vesus HC, pcME/CFS versus nPCS, pcME/CFS versus HC, and nPCS versus HC. Volcano plots were generated to visualize these differential expression patterns, highlighting cytokines with statistically significant fold changes, defined as an absolute log-fold change greater than one, with a significance level set at 10%, due to the exploratory nature of the study.

**Correlation analysis** was performed to examine pairwise relationships between cytokines, as well as between cytokines and clinical scores, as well as anti SARS-CoV2-Nukleocapsid (NCAP) IgG and anti SRAS-CoV2-Spike Protein IgG utilizing Spearman’s rank correlation coefficient. This non-parametric approach was chosen to capture both linear and non-linear associations, providing insights into co-regulatory mechanisms within the immune network. Correlation matrices were generated separately for pcME/CFS, nPCS, and HC groups, allowing for the identification of unique interaction patterns within each study group. Heatmaps, created by using the ggplot2 package, provided a detailed visual summary of immune interactions in the three groups. Significant correlations are marked with [*] for p ≤ 0.01. The classification of the correlation coefficient effect size is set as follows, values between 0 and 0.2 indicate no or a very small effect, from 0.2 up to 0.5 correspond to a small effect, while values from 0.5 up to 0.8 signify a medium effect. Any value from 0.8 and above represents a strong effect. All statistical analyses and visualizations were conducted in R (version 4.2.1) by using RStudio (version 7.1), leveraging specialized packages tailored for each analysis step.

### Rational defining surrogate cytokine marker

Cytokines appearing in the volcano plots with an absolute log-fold change greater than one and a corresponding p-value below 10%, or those identified through a Kruskal-Wallis test with a p-value below 10%, were selected as relevant candidates for further analysis.

### KEGG Analysis

Kyoto Encyclopedia of Genes and Genomes (KEGG) was implemented to understand disease pathogenesis through its detailed pathway maps and databases. For our study, surrogate cytokines with significant correlations, as shown in **Figure 3**, were used as input for KEGG pathway analysis (https://www.genome.jp/kegg) to identify biologically and clinically relevant pathways.

## Results

### The IgG-induced secretome discovered differences in PCS groups and in response to viral proteins

After stimulation with purified IgG, the cytokine profiles of monocytic U937 cells were analyzed by a cytokine array comprising 80 different proteins. To measure AAB-induced cytokine expression between HC and diseased cohorts, principal component analysis (PCA) was employed to stratify high-dimensional data and to uncover distinct cytokine profiles, which could contribute to group separations. Study groups were matched for a similar age distribution and disease duration (**Table 1**). The BMI was not different among the groups. Our data indicate a distinct clustering pattern across the groups (**Figure 1**). When comparing HC to all PCS patients, including those with and without ME/CFS, a clear separation between the two cohorts was identified (**Figure 1A**). Comparison of nPCS versus pcME/CFS demonstrated partially overlapping clustering (**Figure 1B**), indicating shared immune dysregulation while also capturing subgroup-specific immune patterns. PCS is characterized by an abnormal immune response to SARS-CoV-2. To understand the differences in IgG-induced cytokine profiles concerning specific antibodies against SARS-CoV-2 proteins, IgG-induced cytokines were clustered according to the antibodies directed to SARS-CoV-2-spike and SARS CoV2-SARS-CoV2-NCAP. Here, only significantly upregulated cytokines were selected. Interestingly, SARS-CoV-2-specific antibodies did reveal the strongest correlations with IgG-induced cytokines by applying HC-IgG. As examples, spike IgG antibody levels did positively correlate with GRO-α (Growth-Regulated Oncogene Alpha; 0.72, p = 0.01), IL-7 (Interleukin 7; 0.68, p = 0.01), and IL-5 (Interleukin 5; 0.67, p = 0.02). Additionally, positive correlations with antibody levels against SARS-CoV2-NCAP were shown with IGFBP-4 (Insulin-Like Growth Factor Binding Protein 4; 0.74, p = 0.26), IL-4 (Interleukin 4; 0.74, p = 0.26), and SCF (Stem Cell Factor; 0.74, p = 0.26), suggesting well-regulated immune communication (**Figure 1C**). In contrast, cytokine correlations with SARS-CoV-2 spike IgG were markedly absent in nPCS with only a positive correlation with NT-4 (0.59; p=) and a negative correlation with TGF-β3 (Transforming Growth Factor Beta 3) levels (−0.61, p = 0.04). In the pcME/CFS group (**Figure 1C**), strong negative correlations between spike IgG titers and NAP-2 (−0.90, p = 0.00), MCP-3 (−0.81, p = 0.03), and MIP-3α (−0.75, p = 0.03) levels were detected as well as between SARS-CoV2-NCAP IgG ab levels and MCP-3 levels (−0.81, p = 0.03). These data indicate changes in regulatory mechanisms induced by SARS-CoV-2 and a disturbed immune response compared to HC (**Figure 1C**, nPCS).

**Figure 1.**
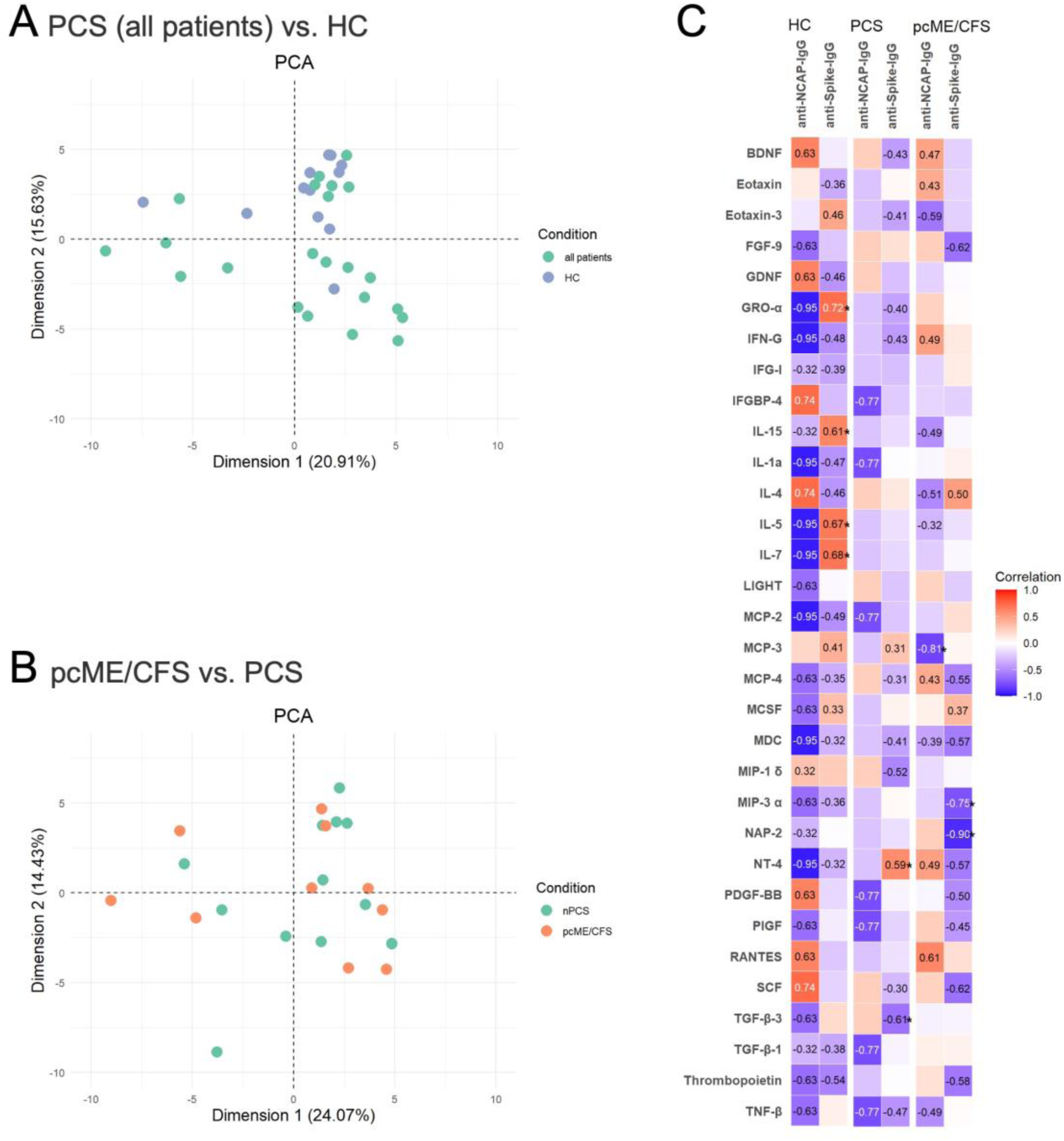
Identification of IgG-induced monocytic secretome in PCS patients compared to HC. PCA analysis indicated that the two cohorts are distinct based on monocytic secretome between PCS patients and HC (A) and between nPCS and pcME/CFS (B). The heatmap revealed an association between spike or NCAP IgGs and secretome factors between nPCS, pcME/CFS, and HC (C).

### nPCS and pcME/CFS AABs mediate distinct cytokine profiles compared to HC

Volcano plots illustrate significant differences in the IgG-induced cytokine expression between patients and HC. Comparing PCS with HC (**Figure 2A**), several cytokines had exhibited significant upregulation such as MIP-1d (Macrophage Inflammatory Protein-1d), PDGF-BB (Platelet-Derived Growth Factor-BB), TGF-β3, and SCF (Stem Cell Factor), all of which had shown both a high log2 fold change and statistically significant p-values. These cytokines are involved in inflammatory, fibrotic, and tissue repair pathways, suggesting persistent immune activation. Further, in PCS, PlGF (Placental Growth Factor), MCP-2 (Monocyte Chemotactic Protein-2), and IL-4 have been also upregulated as markers for angiogenesis, monocyte recruitment, and Th2-related anti-inflammatory responses. Thrombopoietin, a marker for disruptions in coagulation, was modestly upregulated. The IgG-induced secretome also showed differences between pcME/CFS and nPCS (**Figure 2B**). Specifically, IL-1α (Interleukin-1 alpha), GRO-α (Growth-Regulated Oncogene alpha), and Eotaxin-3 (Eosinophil Chemotactic Protein 3) revealed notable increases in expression in pcME/CFS. The detailed comparison between nPCS or pcME/CFS with HC exhibited minor differences in the expression patterns (**Figure 2C** and **D**). MIP-1δ, IL-4 and TGF-β3, revealed comparable upregulation in nPCS as well in pcME/CFS groups compared to HC, suggesting shared features in PCS regardless of ME/CFS status. Interestingly, in the nPCS group compared to HC, a downregulated cytokine levels of GRO-α, IL-5 and IL-7 were found (**Figure 2C**).

**Figure 2.**
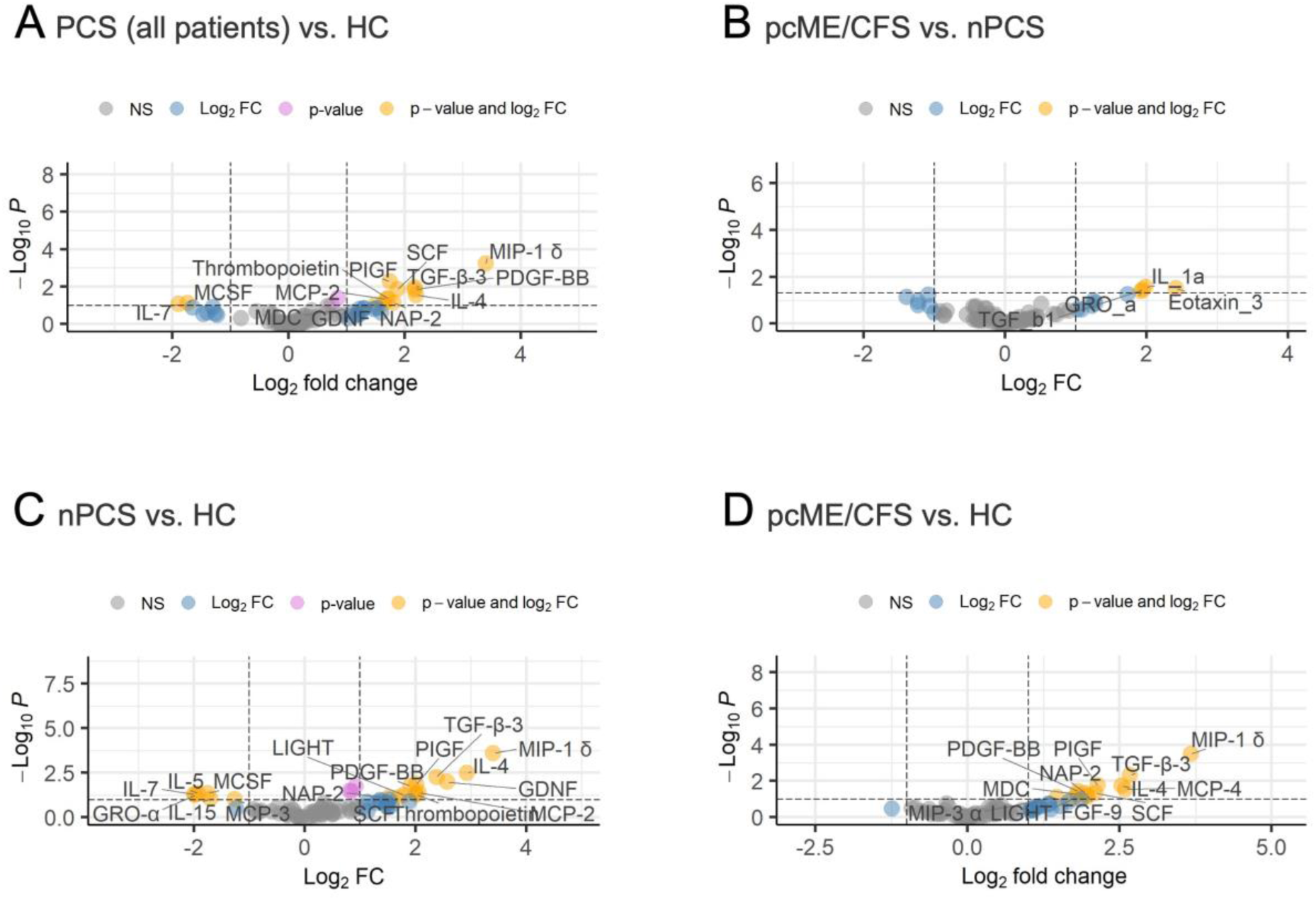
Exploration of cytokine expression between different groups: pcME/CFS&nPCS vs. HC (A), pcME/CFS vs. PCS (B), nPCS vs. HC (C), and pcME/CFS vs. HC (D). The volcano plot illustrates differences in cytokine expression between the groups after stimulation of monocytic cells with IgG.

### AAB-induced secretome identified distinct correlations between cytokines and pathways among the different groups

Intra-cytokine correlations showed marked differences as illustrated in **Figure 3**. In HC, only a few strong correlations such as between FGF-9 and MDC levels (r = 0.90; p<0.001), between Gro-a and IL-5 as well as IFN-g and TGF-b1 (both r = 0.84; p<0.001), (**Figure 3A**). Pathway analysis for HC of the AAB-induced monocyte secretome (**Figure 3B**) revealed a minor number of cytokines involved in autoimmune or infectious disease-related pathways, underscoring the homeostatic nature of HC cytokine interactions. In nPCS (**Figure 3C**), AAB-induced monocyte secretomes reflected altered immune signaling, with strong positive correlations between cytokine levels of IL-15 and IL-5 (r 0.99 p<0.001), GDNF and IGF-I (r = 0.94; p<0.001), IL-15 and IL-7 (r = 0.94; p<0.001) and NAP-2 and PDGF-BB (r = 0.90; p<0.001). Pathways associated with viral infections and rheumatoid arthritis were found, alongside IL-17 signaling pathways. These pathways suggest an overlap between pcME/CFS immune dysregulation and inflammatory responses commonly observed in viral and autoimmune conditions (**Figure 3D**). In pcME/CFS (**Figure 3E**), unique cytokine correlations were found between IL-15 and TNF-b (r = 0.99; p<0.001), IFN-g and TGF-b1 (r = 0.97; p<0.001), FGF-9 and PIGF (r = 0.95; p<0.001) and FGF-9 and MIP-3a (r = 0.91; p<0.001). Pathway analysis of these cytokine networks (**Figure 3F**) revealed strong functional enrichments in pathways associated with rheumatoid arthritis and inflammatory bowel disease, cytokine signaling in viral infection. These findings suggest that pcME/CFS-associated AABs drive immune networks with significant overlap to chronic inflammatory and viral infection-related pathways. In both diseased groups, strong correlations were found between IL-5 and IL-7 (nPCS, 0.95; p<0.001 / pcME/CFS, r = 0.99; p<0.001). Also, moderate correlations in both groups were observed between BDNF (Brain-Derived Neurotrophic Factor) and LIGHT, (nPCS, r = 0.86; p<0.001; pcME/CFS r = 0.88; p<0.001). Noteworthy, in both PCS groups (**Figure 3D, F**) significant fold enrichments were found for RAS-signaling pathways. Cytokine-receptor interactions were found in all groups resembling the nature of cytokine data submitted to KEGG analysis. In sum, the data underscore the critical role of IgG-induced monocyte secretomes in driving disease-specific cytokine interactions and signaling networks.

**Figure 3.**
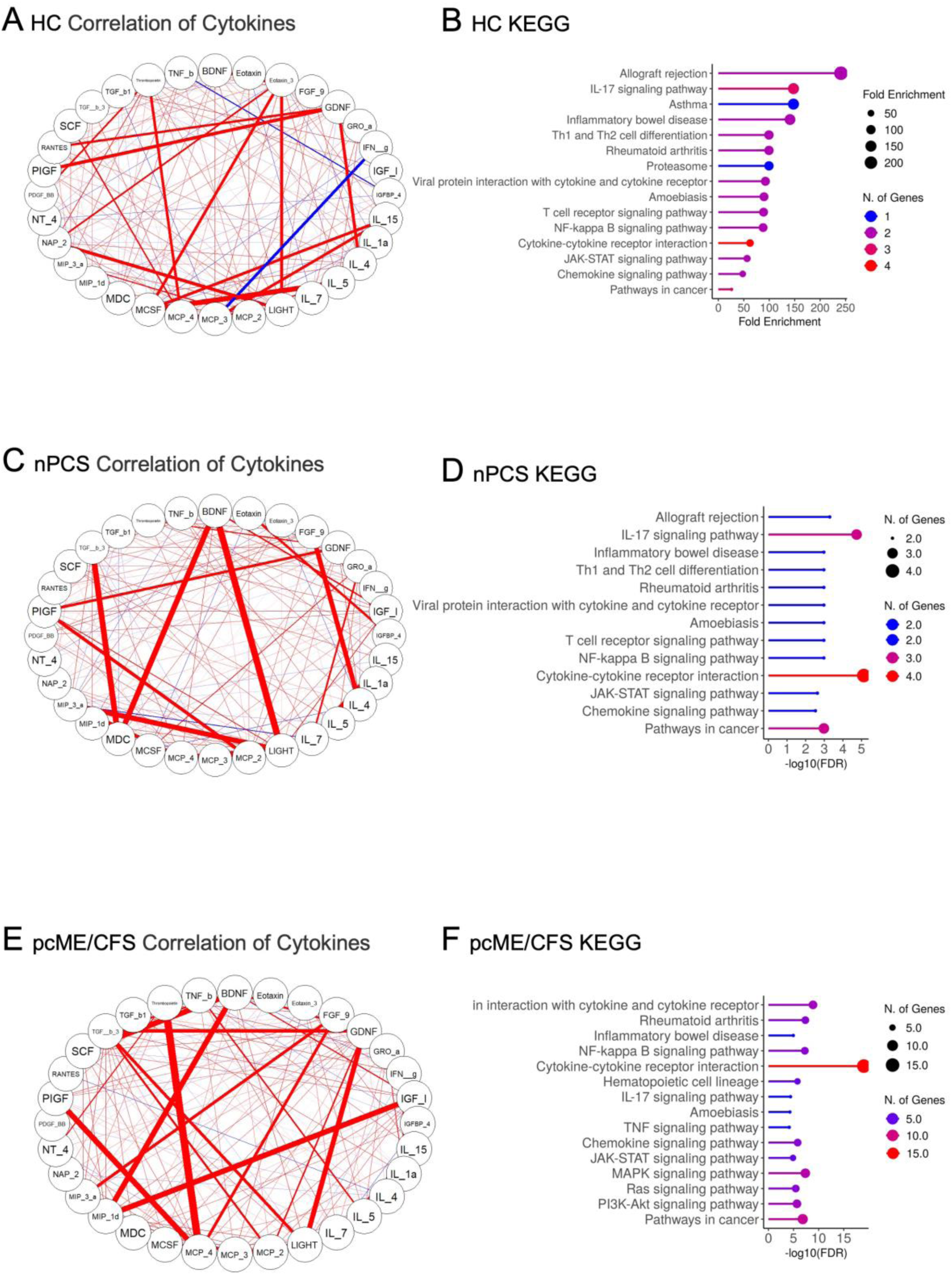
Correlations between cytokines and major signaling pathways in AAB-induced monocyte secretome in HC (A), PCS (B), and pcME/CFS (C). The nodes in the graphs (A, C, and E) represent variables (each cytokine), and a line between two nodes indicates Spearman’s rank correlation coefficient. The line width indicates the strength of the association, with stronger correlations indicated by thicker lines. Multiple connections of nodes indicate clustering between the variables. The bar plot (B, D, and F) indicates the 10 most representative altered canonical KEGG pathways affected by the differentially expressed cytokines. The most significantly enriched pathways are depicted in a progressively redder colour.

### IgG-induced proteins correlate with distinct manifestations in nPCS and pcME/CFS

To explore how IgG-driven cytokine responses may contribute to specific symptoms, IgG-induced protein levels were compared with clinical data (**Figure 4**). In nPCS (**Figure 4 A**), moderate positive correlations were found between cytokine levels and symptom severity (Bell score). Autonomic dysfunction assessed by the total Composite Symptom Score (Compass-31 total), correlated significantly with Eotaxin levels (r = 0.60, p = 0.03), indicating its role in exacerbating symptoms and eosinophil recruitment [26]. In more detail, the domain bladder impairment (Compass-31) showed strong correlations with IGFBP-4 levels (r = 0.75, p < 0.001), PlGF (r = 0.69, p = 0.01), and MCP-2 levels (r = 0.63, p = 0.02), reflecting the involvement of neuroprotective mechanisms, vascular inflammation, and monocyte recruitment in nPCS bladder symptoms [27–29]. Remarkably, severity of fatigue were positively correlated with IL-4 levels (r = 0.66, p = 0.01), emphasizing Th2-mediated immune activation [30] as well as with FGF-9 levels (r = 0.72, p = 0.01). Eotaxin levels were moderately correlated with the degree autonomic dysfunction (r = 0.6, p = 0.03). In nPCS, gastrointestinal symptoms (com_gi) were negatively associated with RANTES levels (r = −0.79, p < 0.001) and NAP-2 levels (r = −0.68, p = 0.01), indicating the role of impaired monocyte chemotactic signaling in eosinophil regulation [31].

**Figure 4.**
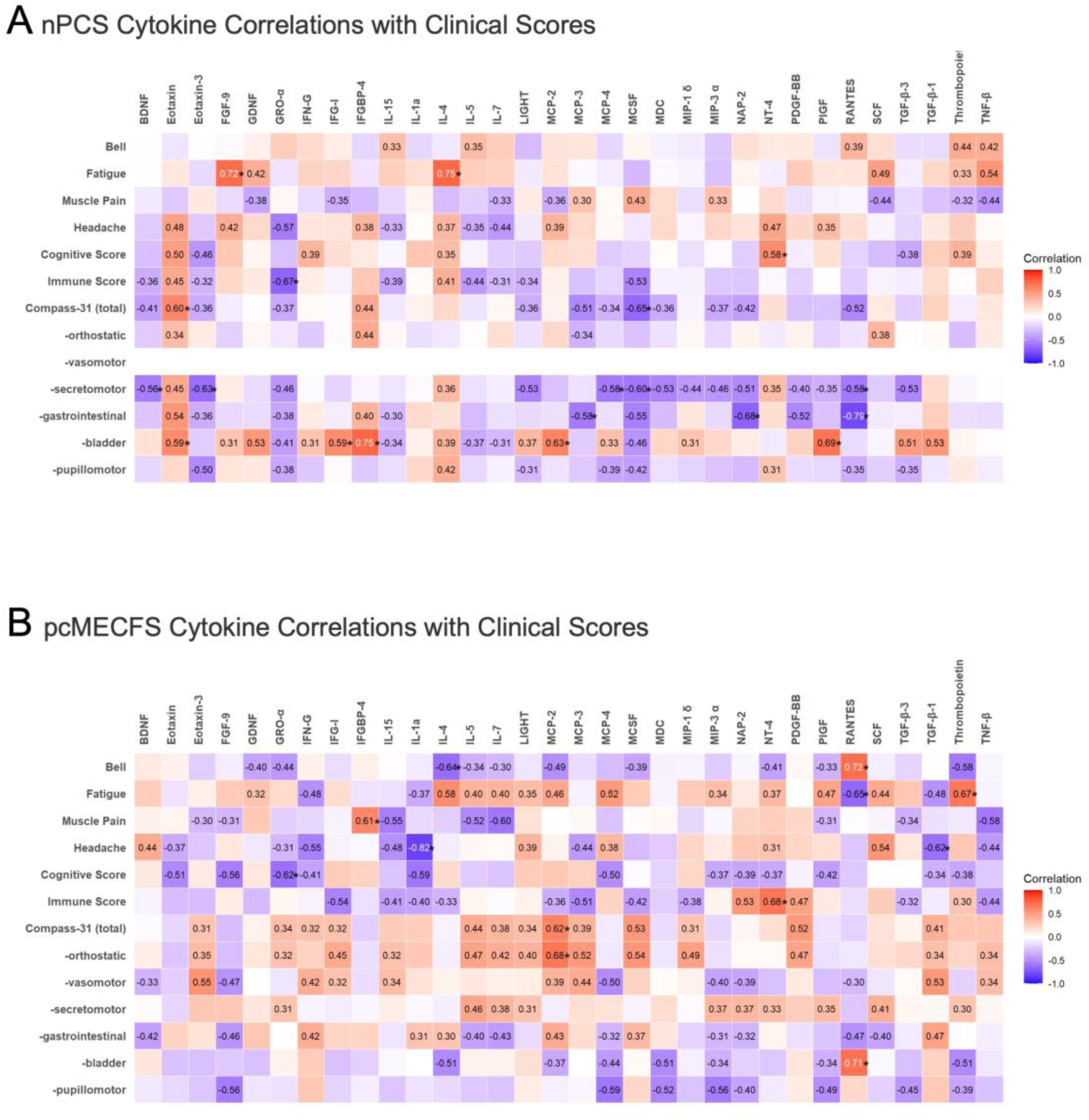
The heat map of the correlation matrix summarizes the results of Spearman correlation analysis among the clinical symptom scores and cytokine data considered in PCS (A) and pcME/CFS (B). Spearman correlation coefficients (r) are indicated in the relative boxes at the intersection between the considered variables. Positive and negative correlation coefficients are shown in red and blue, respectively.

In pcME/CFS (**Figure 4B**), unique cytokine correlations suggested heightened neuroinflammatory and immune dysregulation compared to nPCS. The levels of RANTES, involved in angiogenesis as well as in inflammation, were strongly correlated positively with both the Bell Disability Score (r = 0.72, p = 0.01) and bladder dysfunction (com_blad) (r = 0.71, p = 0.01) [32]. Correlations between the Immune Score and the levels of the neurotrophin NT4 (r = 0.68, p = 0.02) were observed. Interestingly, severity of fatigue was correlated with Thrombopoietin levels (r = 0.62, p = 0.04), implicating platelet activation in fatigue and chronic inflammation [33]. Additionally, levels of IGFBP-4 (r = 0.61, p = 0.05), a neuronal survival factor [27], was correlated positively with the severity of muscle pain. The degree of cognitive dysfunctions was correlated negatively with GRO-α (r = −0.67, p = 0.02), while IL-7 levels negatively correlated with muscle pain severity (r = −0.60, p = 0.05). The levels of the chemokine MCP-2, found increased in PCS patients [29] and causing endothelial impairment [34], was correlated particularly with orthostatic dysfunction (r = 0.68; p = 0.02) and compass-31 (com_total; r = 0.62, p = 0.05). In pcME/CFS, neuroinflammatory markers had shown stronger negative correlations compared to nPCS. IL-1α correlated negatively with severity of headache (r = −0.82, p < 0.001), suggesting its role in neuroinflammation linked to headache symptoms. TGF-β1 also had shown negative correlations with headache severity (r = −0.62, p = 0.04), alterations of regulatory signaling in pcME/CFS [31].

## Discussion

In this study, we aimed to identify common and distinct IgG-driven immune mechanisms, by stimulating monocytes, a supposed key cell in the response to virus infections, with IgG from nPCS and pcME/CFS patients [13, 16, 35]. The observed differences in the responses to viral proteins indicate disturbed immune response already present at the beginning of PCS. In general, PCS patients exhibit distinct patterns of cytokine upregulation associated with immune dysregulation, tissue repair, and chronic inflammation. In addition, key differences between nPCS and pcME/CFS were identified, suggesting that IgG from pcME/CFS patients exhibit more pronounced neurotrophic responses. IgG-induced cytokines exhibit strong correlations with clinical symptoms emphasizing similarities between IgG-induced pathways and those present in the patients. The results of this study strongly indicate a contribution of AAB in the pathogenesis of disease and provide important insights into the immune landscape of PCS. The study highlights potential pathways involved in the persistence of symptoms and the development of chronic fatigue-related conditions.

In pcME/CFS, AABs mediated strong inflammatory and neurotrophic cytokines indicating immune and endothelial dysregulation. The significant upregulation of cytokines such as MIP-1d, PDGF-BB, and TGF-β3 in PCS patients points to persistent immune dysregulation by AABs months after the acute phase of SARS-CoV-2 infection. These proteins are implicated in tissue repair and fibrosis, aligning with recent findings that suggest long-term tissue damage and scarring may be prominent features of PCS [36]. The upregulation of PlGF further supports the hypothesis of ongoing vascular and immune disturbances in PCS patients caused by dysregulation of the autoantibodiome. PlGF has been associated with vascular inflammation and angiogenesis, both of which may contribute to chronic symptoms such as fatigue and shortness of breath observed in PCS patients [8]. The increased levels of IL-4 indicate a mixed immune response, where both pro-inflammatory and regulatory pathways are activated, may reflecting the complex nature of immune recovery in PCS. Moreover, the moderate upregulation of Thrombopoietin may suggest the involvement of platelet activation and coagulation pathways or a negative rebound causing thrombocytopenia, which have been implicated in both acute COVID-19 and long-COVID syndromes [33]. This could contribute to microvascular dysfunction, further exacerbating symptoms such as brain fog and fatigue.

By distinguishing the immune response to IgG between nPCS to pcME/CFS the upregulation of IL-1α, GRO-α, and Eotaxin-3 in pcME/CFS patients aligns with previous studies that have demonstrated immune dysregulation and heightened inflammation in ME/CFS compared to nPCS groups. These cytokines are critical mediators of inflammation, which could contribute to the severe fatigue, post-exertional malaise, and other hallmark symptoms of ME/CFS [37]. pcME/CFS-IgG triggered cytokine profile also reflects a shift toward a more chronic inflammatory state, GRO-α is linked to neutrophil activation and could be contributing to chronic low-grade inflammation, while Eotaxin-3 suggests a potential role for eosinophils, which are involved in allergic responses but also implicated in prolonged inflammatory states [26, 38]. The relatively stable levels of TGF-β1 in this cohort could point to a lack of sufficient regulatory immune responses, which was partly observed in in-vitro experiments with PCS ME/CFS sera, where immune dysfunction persists, additionally resembled by sICAM-1, IGFBPB-4 and cystatin-C [8]. These findings provide crucial insights into the very early AAB-mediated processes occurring in PCS, particularly for patients with ME/CFS. The short-term nature of the experiments enables us to observe immune dysregulation at the onset of the post-viral phase rather than long-term chronic responses.

The results of group specific cytokine correlations highlight distinct cytokine and growth factor interactions in nPCS and pcME/CFS, shedding light on immune mechanisms underlying symptom severity and disease progression. In both groups, while strongest correlations were observed in pcME/CFS, for cytokines such as IL-5, IL-7, IL-15, and TNF-β, reflecting enhanced inflammatory and immune dysregulation. IL-15, associated with disease severity in COVID-19 [39] and TNF-β, a key marker uniquely elevated in ME/CFS after exercise[40], point to chronic immune dysregulation and post-exertional inflammation, characteristic of pcME/CFS. These findings suggest a sustained and amplified immune response compared to pcME/CFS. Growth factors such as FGF-9 and PIGF also exhibited stronger associations in pcME/CFS. FGF-9, documented in movement disorders and ME/CFS, plays a role in tissue repair and is implicated in pulmonary fibrosis [41], while PIGF is linked to angiogenesis and increased after acute exercise [42]. Together with pathway correlations with autoimmune diseases and RAS-signaling, the results indicate angiogenic and fibrotic dysregulations, contributing to tissue remodeling and symptom chronicity in pcME/CFS. These results underscore the importance of cytokines and growth factors induced by patient IgG in both the severity and progression of PCS.

The comparative analysis of the AAB-induced secretome revealed distinct immune pathways underlying symptom manifestations in nPCS and pcME/CFS. These findings emphasize the central role of immune dysregulation in driving fatigue, cognitive dysfunction, and organ-specific symptoms, highlighting the differing contributions of pro-inflammatory and neuroprotective cytokines in these conditions. In nPCS, immune dysregulation was linked to symptom severity, with cytokines such as Eotaxin and IGFBP-4 prominently associated with overall symptom burden and bladder dysfunction, respectively. Eotaxin, involved in eosinophil recruitment and monocyte-driven inflammation, correlated positively with symptom severity, reflecting its role in immune-mediated exacerbation [31]. IGFBP-4, recognized for its neuroprotective properties, showed a strong positive correlation with bladder dysfunction, suggesting a compensatory response to immune-mediated damage in bladder tissues [27]. Thrombopoietin was negatively associated with post-exertional malaise (PEM), linking platelet activation to vascular repair and immune recovery processes [33]. Similarly, RANTES and NAP-2 exhibited negative correlations with gastrointestinal symptoms, reflecting impaired chemotactic signaling and vascular regulation [31, 43]. The pcME/CFS demonstrated heightened immune dysregulation and stronger associations with neuroinflammatory and neuroprotective pathways. RANTES emerged as a dual mediator, positively correlating with bladder dysfunction but better physical function (high Bell score), suggesting its involvement in tissue repair and neuroimmune pathways specific to severe ME/CFS [32]. In fibromyalgia syndrome, where muscle pain and bladder issues coexist, the association of IGFBP-4 with muscle pain points to shared inflammatory pathways [44]. These findings highlight a neuroimmune axis driven by monocytes in response to AABs in both pain and autonomic dysfunction. Noteworthy, bladder function did not correlate with the age of pcME/CFS patients. Neurotrophic factors such as NT4 and PDGF-BB were positively correlated with immune scores, which may indicate compensatory responses to ongoing neuroinflammation and tissue damage, both effectors promote survival of different immune cells [31, 45]. Fatigue in pcME/CFS showed nuanced cytokine interactions. While IL-4 and MCP-2 were positively associated with fatigue, TGF-β1 exhibited a negative correlation, reflecting its protective role in regulating immune-driven fatigue [31, 46]. Intriguingly, MCP-2 upregulation is well described in PCS patients, hypothesizing a key role of AAB-mediated monocytic response in PCS pathogenesis [29]. Neuroinflammation was evident through negative correlations between cognitive function and cytokines such as Eotaxin and GRO-α, emphasizing the impact of chronic inflammation on cognitive impairment [26, 27].

This study demonstrated IgG-induced cytokine signatures in an in vitro assay correlate more pronounced with disease-specific symptoms compared to direct serum AAB measurements or analyses of specific antibodies, such as β2-adrenergic receptor antibodies in ME/CFS [14, 16]. These findings suggest that IgG-induced cytokine responses provide a more accurate assessment of pathogenic potential. An in vitro diagnostic developed in parallel by the Clinic for Rheumatology and Clinical Immunology (University Clinic Schleswig-Holstein; placeholder (EP25150270.4) further validated these results, showing a sensitivity of 92% and specificity of 83% in distinguishing PCS patients from HC, and for the first time differentiating nPCS from PCME/CFS patients. This approach highlights its potential for precise diagnosis and disease prognosis.

## Conclusion

The findings from this study suggest that the immune exhaustion observed in monocytes stimulated by IgG from nPCS and pcME/CFS patients could be driven by chronic activation through dysregulated AABs. This has been observed in other chronic inflammatory conditions, where persistent immune stimulation, like viral infections, lead to immune cell exhaustion and endothelial impairment [47–49]. Thus, AABs may maintain a state of chronic immune activation, which, over time, results in monocyte dysfunction and dedifferentiation, mirroring processes seen in autoimmune diseases and chronic infections [50, 51], subsequently contributing to immune exhaustion and causing long-term symptoms observed in PCS with ME**/**CFS.

## Data Availability

All data produced in the present study are available upon reasonable request to the authors.
All data produced in the present work are contained in the manuscript.

https://www.gesundheitsforschung-bmbf.de/en/index.php

## Acknowledgments

We are grateful to the patients who participated in this study despite their individual disease related impairment.

